# Nasogastric tube in critical care setting: combining ETCO2 and pH measuring to confirm correct placement

**DOI:** 10.1101/2021.06.15.21258970

**Authors:** S. Ceruti, S. Dell’Era, F. Ruggiero, G. Bona, A. Glotta, M. Biggiogero, E. Tasciotti, C. Kronenberg, A. Saporito

## Abstract

**Introduction:** nasogastric tube (NGT) placement is a common procedure performed in critical care setting. Chest X-Ray is the diagnostic gold-standard to confirm correct placement, with the downsides of both the need for critical care patients’ mobilization and intrinsic actinic risk. Other potential methods to confirm NGT placement have shown lower accuracy compared to chest X-ray; ETCO_2_ and pH analysis have singularly yet investigated as an alternative to the gold standard. Aim of this study was to determine thresholds in combine measurements of ETCO_2_ and pH values, at which correct NGT positioning can be confirmed with the highest accuracy.

**Material & Methods:** a prospective, multicenter, observational trial; a continuous cohort of eligible patients was allocated to two arms, to identify clear cut-off threshold able to detect correct NGT tip positioning with the maximal accuracy. Patients underwent general anesthesia and orotracheal intubation; in the first group difference between tracheal and esophageal ETCO_2_ values were assessed. In the second group difference between esophageal and gastric pH values were determined.

**Results:** from November 2020 to March 2021, 85 consecutive patients were enrolled: 40 in the ETCO_2_ group and 45 in the pH group. The ETCO_2_ ROC analysis for predicting NGT tracheal misplacement demonstrate an optimal ETCO_2_ cutoff value of 25.5 mmHg, where both sensitivity than specificity reach 1.0 (AUC 1.0, p < 0.001). The pH ROC analysis for predicting NGT correct gastric placement demonstrated the optimal pH cutoff value at 4.25, with a mild diagnostic accuracy (AUC 0.79, p < 0.001).

**Discussion:** A device capable of combining the presence of a *negative marker* with a *positive marker* could be accurate enough in identifying the correct NGTs positioning. Further studies are required to validate the reproducibility of these results by a specific device, whose accuracy also ought to be compared with standard chest X-ray.

Trial number: NCT03934515 (www.clinicaltrials.gov)

## Introduction

Nasogastric tube (NGT) placement is an extremely common procedure routinely performed on patients in critical care setting.[1] This simple procedure however is not riskless: serious complications can occur, especially in sedated, intubated and paralyzed patients, when cough reflex have been abolished[2]. The incidence of complications during NGT positioning is around 4%, with both a high morbidity[3], possibly leading to a prolonged hospital stay and higher costs and an increased mortality[4].

Currently, the gold standard for correct positioning confirmation is the Chest X-Ray[5], which implies the use of ionizing radiation (4 μSv for radiography). This may not be negligible considering that critical patients, during their hospital stay, may require multiple NGT placements and repositioning after displacement. Furthermore, in critically ill patients the image receptor is placed directly under the back, requiring their passive mobilization, which increases both staff workload and clinical risks (for example of accidental extubation or hemodynamic instability).

Several alternatives have been investigated in order to confirm correct NGT placement; these include the so-called *bubble technique*[6], frozen NGT[7], gastric auscultation[8,9], aspiration from the NGT[10,11], gastric ultrasound[12–14], biochemical markers[15],[16] and the use of magnets[17]; however none of them was associated to a high diagnostic accuracy. Some pilot studies have shown that measurements of end tidal CO_2_ (ETCO_2_) with a graphic capnometer could be used to determine whether NGT tip has been erroneously placed at tracheal level[18–22], others that pH measurements can distinguish a correct NGT tip position from an esophageal positioning, by analyzing the relative pH levels[23,24]. In those studies, however, threshold values of ETCO_2_ and pH able to discriminate between correct and failed positioning have not been determined: the two measurements had never been associated, the global accuracy of each methodology was rather low, or the sample size was insufficient to obtain a statistical significance. The feasibility to use these two parameters in a hypothetical device exploiting a double feedback mechanism to detect correct NGT placement with a high accuracy is an attractive possibility which demands further investigations.

Aim of the study is to analyze distributions between tracheal and esophageal ETCO_2_ values and between gastric and esophageal pH values, in order to identify thresholds values at which the correct positioning of NGT can be confirmed with high accuracy.

## Material & methods

This is a prospective, multicenter, observational trial, conducted in a period of six months in two different acute Hospitals. The study has been registered (clinicaltrials.gov, NCT03934515) and approved by the regional Ethical Committee (Comitato Etico Cantonale, Bellinzona, Switzerland, Chairman Prof. Zanini – N. CE3548). A consecutive cohort of patients undergoing general anesthesia, with orotracheal intubation were enrolled; inclusion criteria were patients of both sexes. aged more than 18 years, fasting for at least six hours, who underwent general anesthesia and orotracheal intubation, for whom an oro/naso-tracheal tube positioning was planned according to clinical criteria. Exclusion criteria were patient’s refusal or inability to give informed consent. pregnancy, known ongoing gastric or esophageal bleeding, coagulation impairment (defined as thrombocytes < 50 G/L, fibrinogen < 1.0 g/L, INR > 2.5, aPTT > 70 sec tested at the preoperative assessment), history of traumatic brain injury or polytrauma, esophago-tracheal fistulas, esophageal varices, ENT malformations and/or tumors, history of radiotherapy for ENT tumors. Patients in whom pH and/or ETCO_2_ values were not measurable for technical reasons were excluded and considered as drop-outs.

### Randomization and allocation

Eligible patients were allocated 1:1 to one of the two groups. Group A underwent ETCO_2_ measurement; group B patients underwent pH measurement.

### Group A

After anesthesia induction and orotracheal intubation, a suction probe was inserted within the endotracheal tube and tracheal ETCO_2_ were measured at 15 seconds through a capnometer connected to the probe. Both the probe and the NGT had the same diameter (12 Fr). After the measurement, secretions were aspirated as usual. Then, NGT was positioned using the standard approach and esophageal ETCO_2_ was measured at 15 seconds through a capnometer attached to the NGT. After the measurement, the capnometer was disconnected, while the NGT was left in palace as usual. ETCO_2_ values were registered in the data sheet and subsequently transferred into an anonymized electronic database; two sets of values were therefore obtained for each patient: tracheal and esophageal ETCO_2_.

### Group B

After anesthesia induction and orotracheal intubation, NGT insertion was performed according to local protocols. pH was than measured using litmus paper on liquid aspirated from the NGT. NGT was progressively inserted, measurements were taken on secretions aspirated from NGT at two different steps at two insertion depths: 25 cm from teeth (esophageal level) and at 40 cm from teeth (gastric level). If it was not possible to aspirate some fluid, 10 ml of NaCl 0.9% were inserted and then aspired back and pH measurements were taken on this washing fluid. All values were registered and archived as described above; two sets of values were therefore obtained for each patient: esophageal and gastric pH.

### Statistical analysis

According to literature, a tracheal ETCO_2_ value around 40 mmHg[25–27] was assumed as normal, while a normal esophageal ETCO_2_ was around 20 mmHg [28]. In order to have a significant difference between tracheal and esophageal groups, with a power of 90% and a significance level of 0.01 (one-tailed z test), we calculated a numerosity of 35 patients. Anticipating a 10% drop-out rate, we included 40 patients for group A. With regard to the pH arm, we assumed an esophageal pH level normal value around 7[29] and gastric pH level ranging from 1.0 to 2.5 [30]. In order to have a significant difference between esophageal and gastric pH values, with a power of 90% and a significance level of 0.01 (one-tailed z test), we calculated a numerosity of 30 patients. Anticipating a 10% of drop-out rate, we included 35 patients for each measurement in group B. We tabulated the distribution of baseline variables across the study’s sections, summarizing categorical variables by frequencies and percentage and numerical variables either by mean and standard deviations (±SDs), or by medians and interquartile ranges (IQR). Data distribution was verified using a Kolmogorov-Smirnov test. We executed a z-test for comparison of the two proportions, refusing the null hypothesis of no difference between the two if the p-value was ≤ 0.01. In order to identify the threshold value of ETCO_2_ which signaled endotracheal positioning of the NGT and the pH value threshold who identified a correct gastric location of NGT with high accuracy, the area under the receiver operating characteristic (ROC) curve was calculated for both ETCO_2_ and for pH level, calculating the sensitivity, specificity and the likelihood ratios for the optimal cut-off point (CP) of the scale (Youden index and Number Necessary to Diagnose, *J* and *NND* respectively)[31]. Starting from the ROC curve, a “cumulative distribution analysis” (CDA) was performed[32], to better identify the gray zone starting from the zone defined by the values associated with a sensitivity and a specificity both of 90%[33]. All hypothesis tests were one-tailed and considered significant if p-value was ≤0.01. Statistical analysis was performed using SPSS.26 (IBM, Chicago, IL, USA) for MacOS.

## Results

From November 2020 to March 2021, 85 consecutive patients were enrolled: 40 in the ETCO_2_ group and 45 in the pH group. 19 dropouts occurred, due to incomplete information sampling during the procedure (such as the impossibility to measure pH); 68 patients were therefore included in the analysis, 33 in the ETCO_2_ group and 35 in the pH group. Mean age was 54 years old (min/max 46 – 62) and 36 (55%) were men. All demographic data are reported in Table 1.

**Table 1.** Demographics characteristic population. Demographic characteristics and main study measurements. Data distribution were expressed as mean ± SD (min-max) or median (25^th^ – 75^th^) if they are not normally distributed according to Kolmogorov-Smirnov test.

|  | ETCO<sub>2</sub> group | pH group | p value |
| --- | --- | --- | --- |
| Age | 54 (13.7) | 60 (9.8) | 0.09 |
| Sex male | 22 (66%) | 14 (43%) |  |
| BMI [Kg/m<sup>2</sup>] | 25.4 (6) | 30.6 (7.4) | 0.36 |
| Systolic arterial pressure [mmHg] | 145 (29) | 152 (28) | 0.36 |
| Heart rate [bpm] | 80 (21) | 77 (17) | 0.43 |
| Respiratory rate [min] | 14 (3) | 14 (2) | 0.7 |
| Jatal hernia | - | 6 (18%) | - |
| PPI intake | - | 12 (36%) | - |
| Esophageal Reflux disease | - | 13 (39%) | - |
| COPD | 7 (21%) | - | - |
| Pulmonary Embolism | 1 (3) | - | - |
| Heart disease | 5 (15) | - | - |
| Tracheal value | 40 (7.1) | - | - |
| Esophageal value | 11 (9.3) | 5.1 (1.3) | - |
| Gastric value | - | 3.2 (1.7) | - |

### Distribution analysis

With regard to the ETCO_2_ *distribution analysis*, 22 (66%) patients were men, 7 (21%) presented a diagnosis of COPD (4 patients of second degree, 1 patient of third degree); one (3%) patient presented a previous diagnosis of pulmonary embolism. Five (15%) patients had a history of heart disease (two patients with severity NYHA 1, three patients with NYHA 2) (Table 1), all with an EF greater than 50%. Mean tracheal ETCO_2_ was 40 mmHg (SD 7.14), while mean esophageal ETCO_2_ resulted 11 mmHg (SD 9.3); a t-test score (Fig. 1) confirmed a significant difference (CI 99%, 24-33, p < 0.001).

**Fig. 1.**
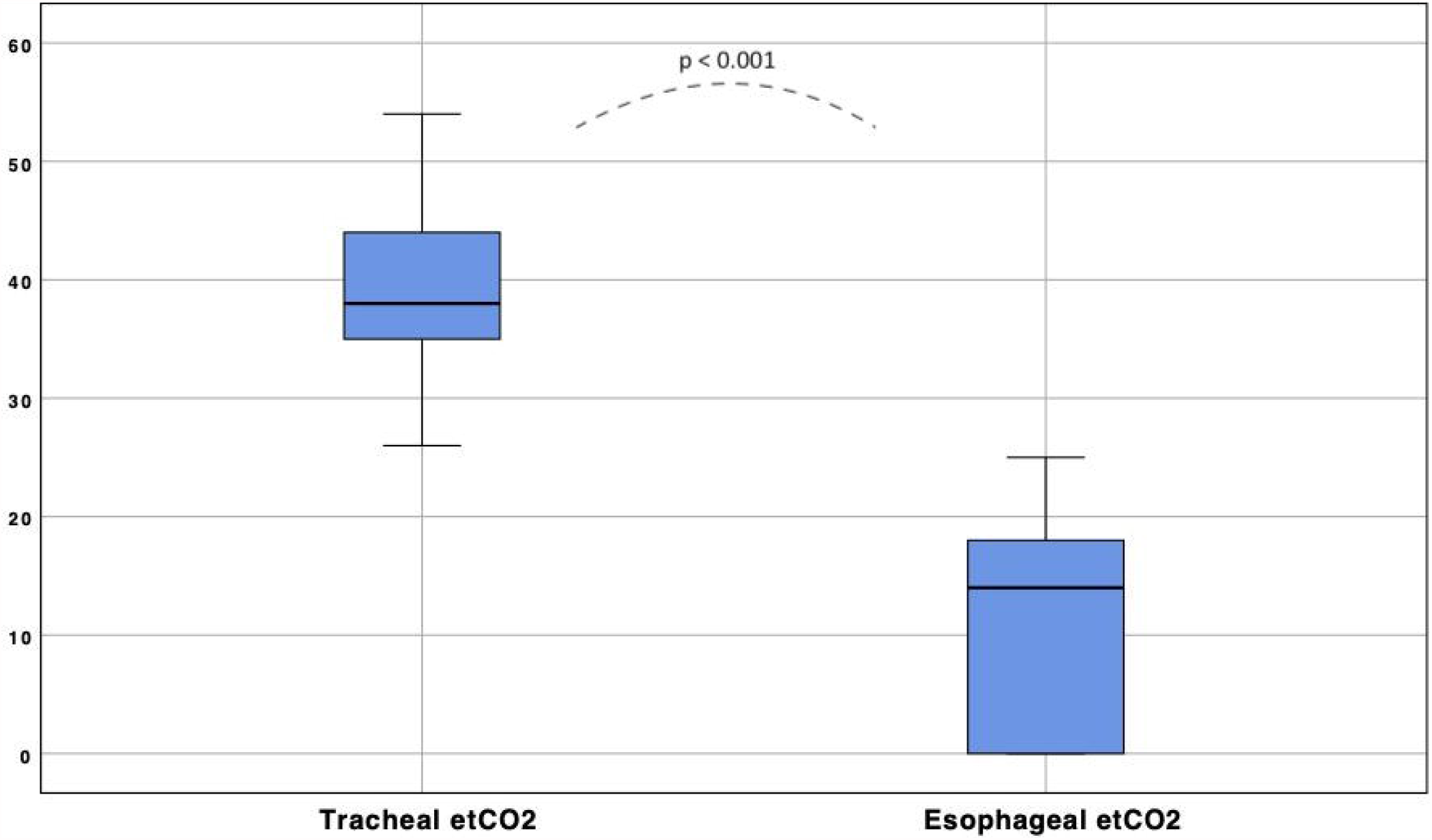
Tracheal and esophageal ETCO_2_ distribution. Boxplots: the black bar indicates median ETCO_2_ (38 mmHg and 14 mmHg respectively), while the blue areas include the interquartile ranges for each group.

Regarding *pH distribution analysis*, 14 (40%) patients were male, 6 (18%) presented a history of hiatal hernia, and 13 (39%) presented a diagnosis of gastroesophageal reflux disease, with 12 (36%) patients receiving PPI therapy at the time of data sampling (Table 1); no patient was on enteral feeding during the analysis. Median gastric pH was 3.1 (1.6 – 4.95), while median esophageal pH resulted 5.15 (4.52 – 6.0); a t-test score confirmed a significant difference (CI 99%, 0.9-2.9, p = 0.004, Fig. 2).

**Fig. 2.**
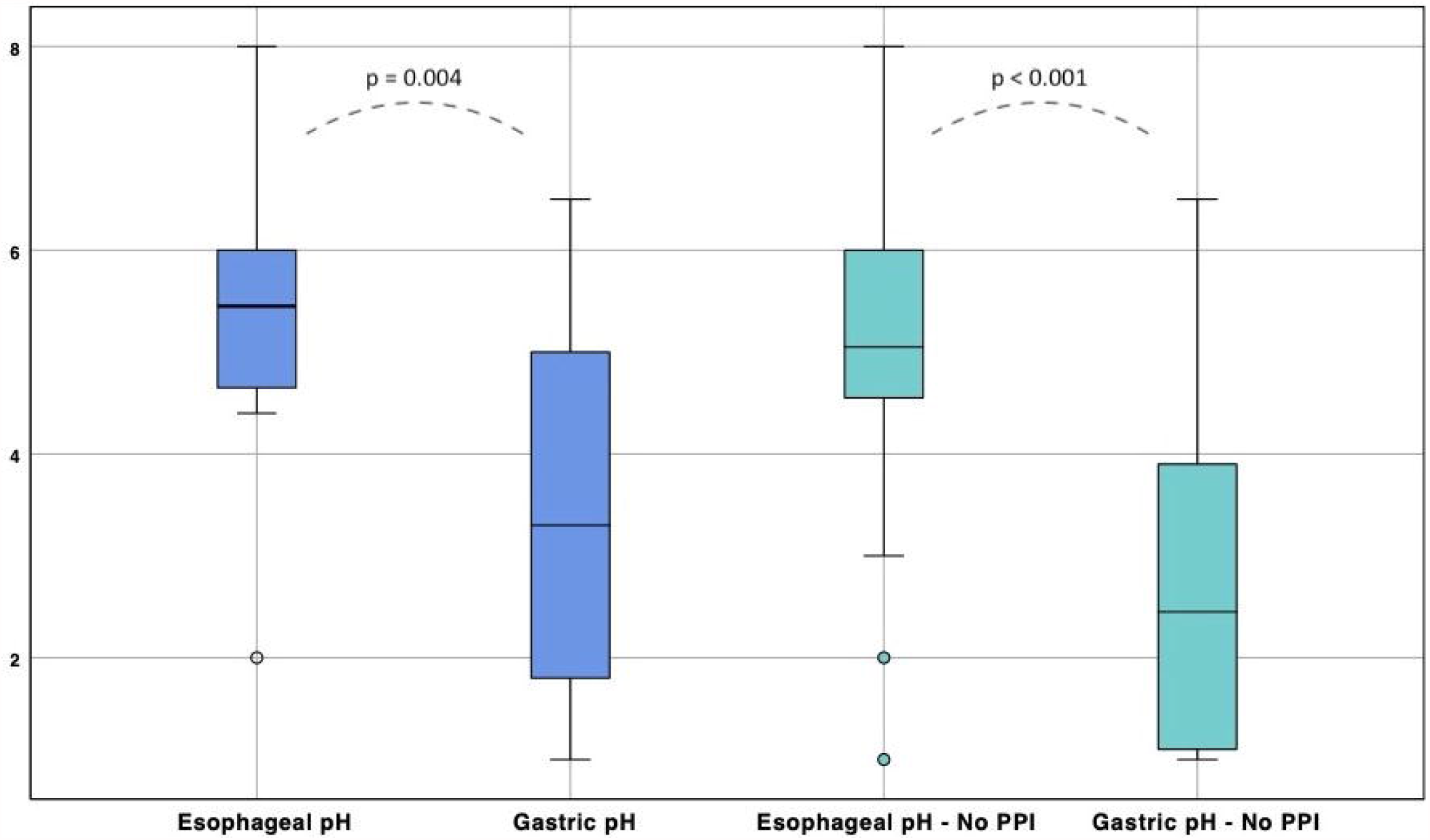
Measured of esophageal and gastric pH. Boxplot distribution in all patients and in patients without PPI use. Regarding the whole group analysis, a t-test score confirmed a significant difference between esophageal and gastric values (CI 99%, 0.9-2.9, p = 0.004). The subgroup analysis involving patients without PPI showed a greater difference (CI 99%, 1.2 – 3.1, p < 0.001) compared the whole group. The black bar indicates median pH, while the blue areas include the interquartile ranges for each group.

A subgroup analysis involving 20 (62.5%) patients without PPI, showed a median gastric pH of 2.45 (1.05 – 4.05) and a median esophageal pH of 5.05 (4.52 – 6.0), with a greater difference of t-test score (CI 99%, 1.2 – 3.1, p < 0.001) compared to all patients (Fig. 2). A comparison between the mean esophageal pH value in all patients with the mean esophageal value in patients without PPI resulted not significantly different (5.1 vs 4.9, p = 0.265).

### ROC curve analysis

The ETCO_2_ ROC curve analysis for predicting NGT tracheal misplacement (Fig. 3A) demonstrate a perfect diagnostic accuracy with an AUC of 1.0 (CI 95%, 1.0 to 1.0, p < 0.001); the optimal cutoff value resulted in an ETCO_2_ value greater than 25.5 mmHg (Youden index J = 1), where both sensitivity than specificity reach 1.0. The pH ROC curve analysis for predicting NGT correct gastric placement (Fig. 3B) demonstrated a mild diagnostic accuracy with an AUC of 0.79 (CI 95%, 0.67 to 0.90, p < 0.001); the optimal cutoff value was a pH below 4.25 (Youden index J = 0.593), with a sensitivity of 0.908 and a specificity of 0.687.

**Fig. 3.**
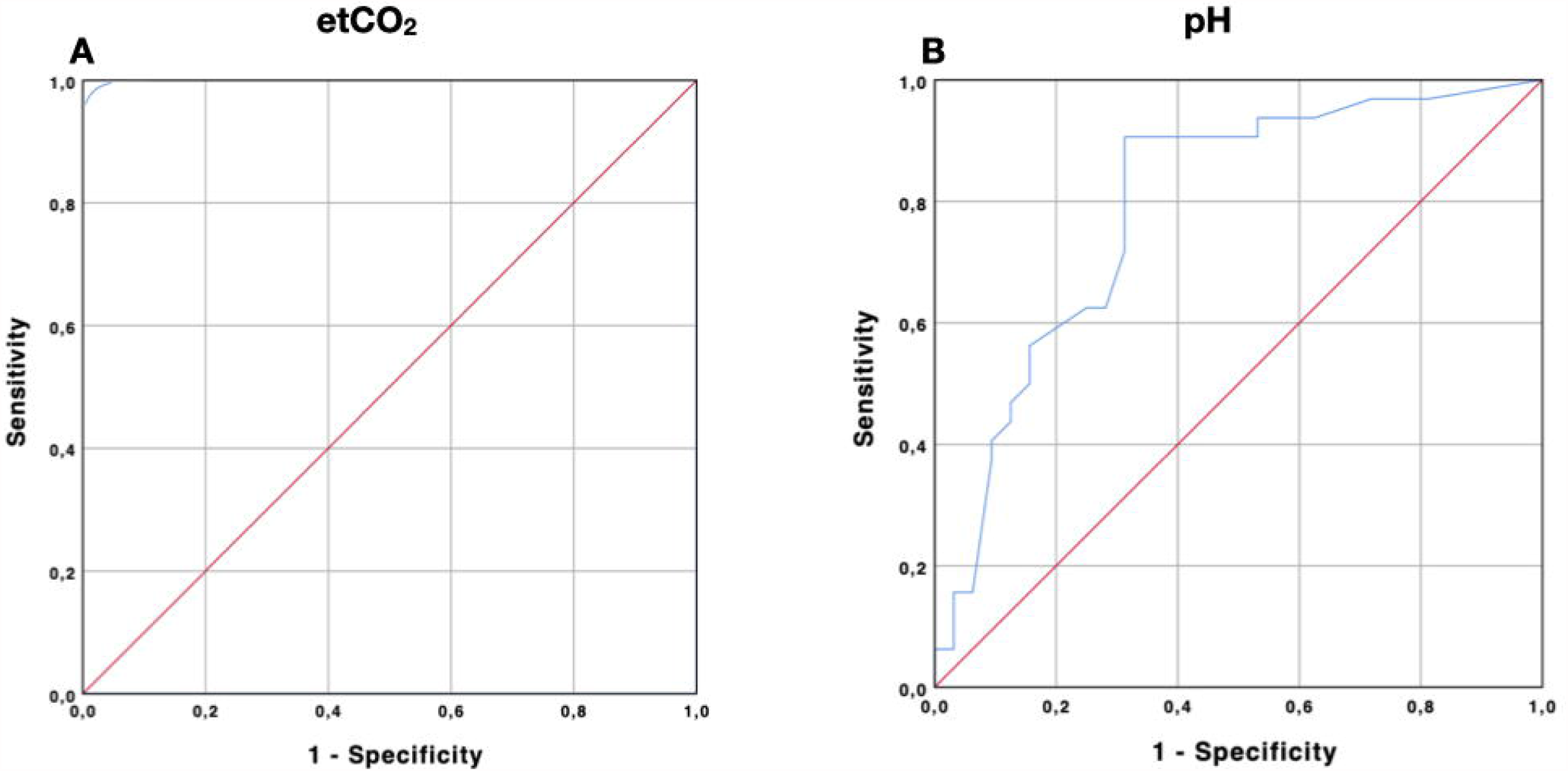
ROC curves of EtCO_2_ and pH method. Receiver operating characteristic (ROC) curves showing the ability of the EtCO_2_ method (figure 3A) and pH method (Figure 3B) to respectively identify a tracheal misplacement (ROC AUC 1.0, p < 0.001) or a gastric correct placement (ROC AUC 0.79, CI 95% 0.67 – 0.90, p < 0.001).

The subgroup analysis involving only patient without PPI confirmed a mild diagnostic accuracy, with an AUC of 0.78 (CI 95%, 0.63 – 0.93, p = 0.002) with an optimal cutoff pH value below 3.9 (Youden index J = 0.6). The NND obtained for the ETCO_2_ method resulted 1, while the NND for the pH method was 1.68 (1.66 in patients without PPI).

Gray zone plots were drawn throughout CDA curves starting from Youden index (Fig. 4) between the 90% of sensibility and 90% of specificity on the two sigma curves for each method (ETCO_2_ and pH); for the pH the gray zone lay between 4.25 and 5.7 (Fig. 4) while for the ETCO_2_ no gray zone was identified, as the tracheal and the esophageal distribution did not cross each other (Fig. 5).

**Fig. 4.**
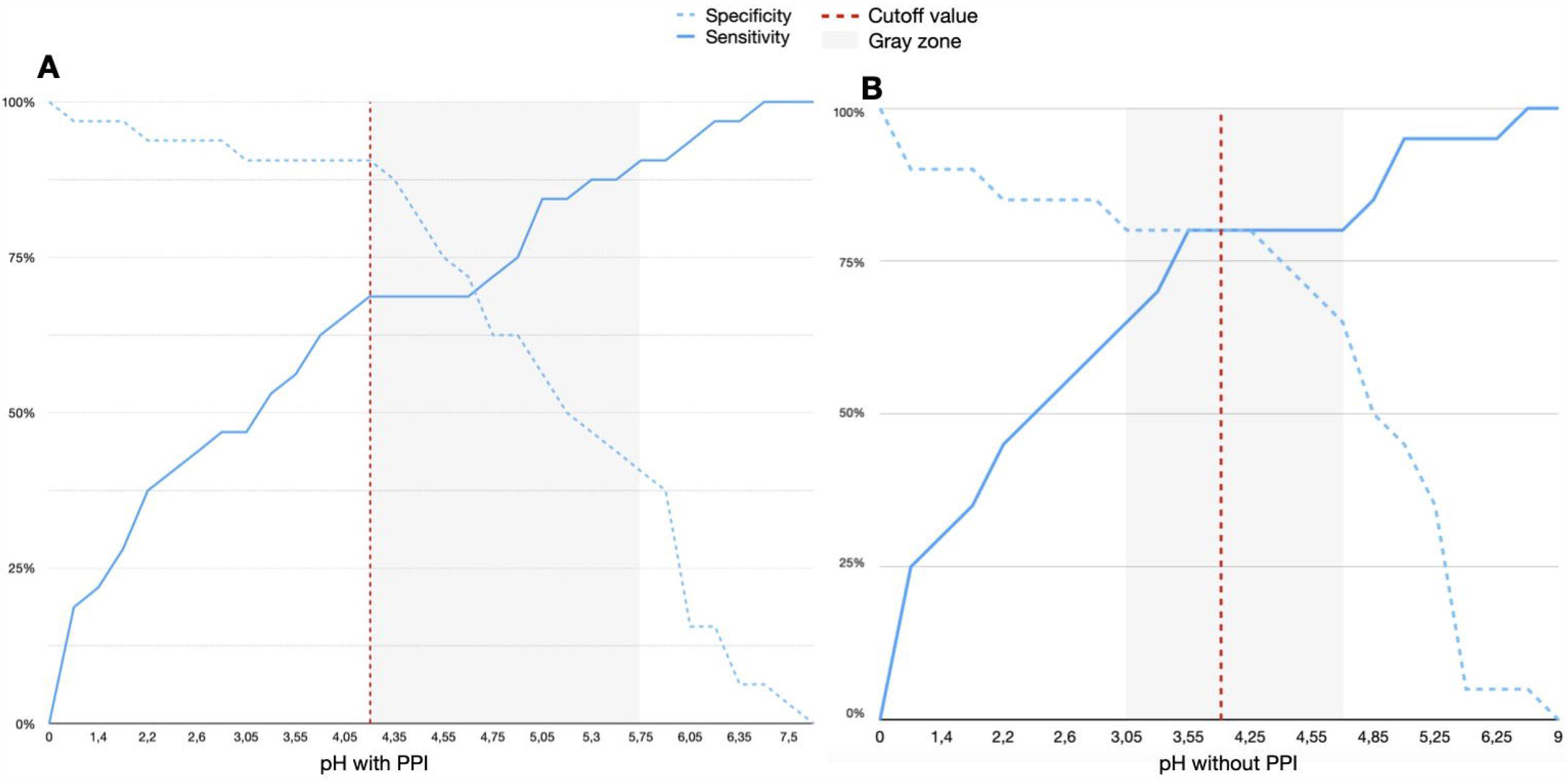
*Cumulative Distribution Analysis* of pH detection. Performed to determine the correct NGT placement with (4A) and without (4B) PPI use. The red line indicates the cutoff limit according to Youden Index (pH below 4.25 and pH below 3.9, with J = 0.593 and J = 0.6 respectively); the gray zone is shown, according with a sensibility than a specificity of 90%.

**Fig. 5.**
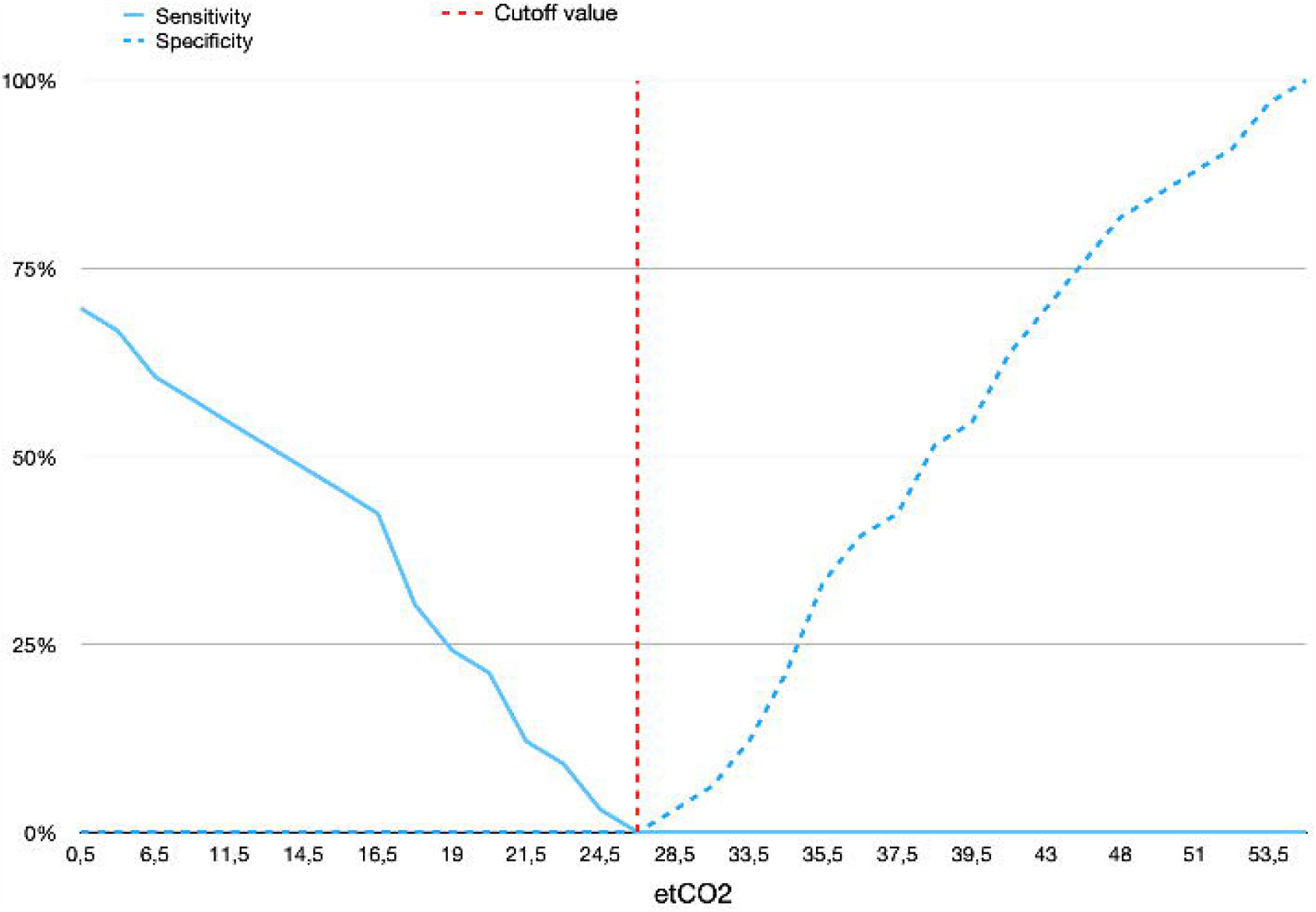
*Cumulative Distribution Analysis* of EtCO_2_ detection. Performed to exclude the NGT misplacement. The red line indicates the cutoff limit according to Youden Index (J = 1).

## Discussion

Nasogastric tube placement in sedated and intubated patients is a procedure potentially associated with dangerous complications. The gold standard to assess correct positioning is Chest X-Ray, which exposes patients to mobilization-related complications, such as devices displacement and hemodynamic and respiratory instability, as well as to actinic risk, due to the need of multiple X-rays.

Alternatives, such as pH and ETCO_2_ measurements, when singularly analyzed, failed to show a superiority compared to chest X-Ray in determining correct NGT tip position[18,23,34], in particular due to lack of identification of an actual threshold value. Our study combines these two techniques, in order to investigate if, when applied together, they can accurately detect a correct positioning of the NGT tip. The use of a double feedback mechanism could in fact prove more accurate than one single technique. In this study, ETCO_2_ distribution between the trachea and esophagus was evaluated intended as a potential *negative marker* to detect NGT misplacement in the upper airways; at the same time, pH distribution between stomach and esophagus was evaluated as a potential *positive marker* for NGT correct placement. Significant differences between tracheal and esophageal ETCO_2_ measurements allowed a complete differentiation in the curve plotting distribution. Based on these results, the use of a qualitative capnometer connected to the NGT and set to detect the threshold value of 25.5 mmHg would be a potentially accurate *negative-marker* mechanism for tracheal NGT placement, with a very high sensitivity, thus avoiding any NGT misplacement.

Concerning the differences in results between gastric and esophageal pH, the distributional differences between the two obtained curves is not neat, especially in case of proton pump inhibitors usage, although extremely low pH values were shown to have a high specificity for gastric NGT placement. Fernandez et al published a review of diagnostic studies to test pH of aspirate fluids using a litmus paper; with this method, they evaluated if the NGT had been correctly positioned. It is to be noted that litmus paper color variation could report a value lower than the actual gastric pH, due to the litmus paper insufficient sensitivity[16]. A recent clinical trial by Gilbertson et al identified the cut off pH < 5.5 to distinguish between the correct positioning in the stomach[23]. In comparison with these two studies, in our trial, the threshold pH value able to minimize false positive rate, thus increasing the specificity of this *positive marker*, resulted 4.25. Noticeably, even if specificity for very low pH appears to be high, the oppositely low consequent sensitivity would affect the global test accuracy thus invalidating the *positive marker* mechanism for detection of gastric NGT placement (NND = 1.68), leading to potential misses of correct placement.

Furthermore, analyzing the data based on PPI therapy allows determining an even lower pH threshold for patients not receiving this class of medications (pH of 3.9), however guaranteeing the same accuracy. In practice, a pH threshold of 4.25 would therefore assure an even better specificity in this subgroup of patients.

Based on our study, a device capable of combining the presence of a *negative marker* (such as ETCO_2_) with a *positive marker* (such as pH) could be accurate enough in identifying the correct positioning of NGTs. Further studies are required in order to validate the reproducibility of these results by a specific device, whose accuracy also ought to be compared with chest X-ray, the current diagnostic gold standard.

This study presented some limitations. This was a preliminary study assessing determined physiological variables; it is still unknown whether a device simultaneously sensing ETCO_2_ and pH could determine correct NGT placement with high accuracy. The presumed esophageal and gastric NGTs placement have been determined based on the distance of the NGT tip from the teeth; there is not, therefore, complete certainty about NGT tip location. However, NGT placed in esophageal site a 40 cm was considered to be placed into the stomach. Moreover, the accuracy of the pH threshold value for the discrimination between esophageal and gastric NGT positioning resulted suboptimal. The use of a normal saline injection in order to measure pH on the aspirated fluid in case secretions could not be aspirated may have affected pH values in these cases.

## Conclusions

In a critical care setting, the use of a device capable of combine the presence of a negative marker to exclude NGT misplacement (like ETCO_2_) and a positive marker to confirm correct NGT placement (as pH evaluation) could be quite accurate to improve correct NGT placement in unconscious patients. Further studies in this direction are needed in order to test this hypothesis.

## Data Availability

Data are available under specific request

## Notes

### Competing Interest Statement

The authors have declared no competing interest.

### Clinical Trial

NCT03934515

### Funding Statement

No funding was obtained for this study.

### Author Declarations

Comitato Etico Cantonale Chairman Prof. Zanini, N. CE3548 Via Orico 4 6500, Bellinzona

